# Towards a brain signature of chronic pain using cerebral blood flow spatial covariance analysis in people with chronic knee pain

**DOI:** 10.1101/19008326

**Authors:** Sarina J. Iwabuchi, Yue Xing, William J. Cottam, Marianne M. Drabek, Arman Tadjibaev, Gwen S. Fernandes, Kristian K Petersen, Lars Arendt-Nielsen, Thomas Graven-Nielsen, Ana M. Valdes, Weiya Zhang, Michael Doherty, David Walsh, Dorothee P. Auer

**Affiliations:** Versus Arthritis Pain Centre, University of Nottingham, Nottingham, UK; NIHR Nottingham Biomedical Research Centre, Queen’s Medical Centre, University of Nottingham, Nottingham, UK; Sir Peter Mansfield Imaging Centre, School of Medicine, University of Nottingham, Nottingham, UK; Division of Rheumatology, Orthopaedics and Dermatology, School of Medicine, University of Nottingham, Nottingham City Hospital, Nottingham, UK; SMI, Department of Health Science and Technology, Faculty of Medicine, Aalborg University, Aalborg, Denmark; Center for Neuroplasticity and Pain (CNAP), SMI, Faculty of Medicine, Aalborg University, Aalborg, Denmark

**Author notes:** Correspondence to: Professor Dorothee P. Auer, Radiological Sciences, Room W/B 1441, Queen’s Medical Centre, Nottingham, NG7 2UH, UK.

**Keywords:** Chronic Pain, ASL, cerebral blood flow, PCA, Knee Osteoarthritis, MRI, experimental pain, sensitisation

## Abstract

Chronic musculoskeletal pain is a common problem globally. Current evidence suggests that maladaptive modulation of central pain pathways is associated with pain chronicity following e.g. chronic post-operative pain after knee replacement. Other factors such as low mood, anxiety and tendency to catastrophize seem to also be important contributors. We aimed to identify a chronic pain brain signature that discriminates chronic pain from pain-free conditions using cerebral blood flow (CBF) measures, and explore how this signature relates to the chronic pain experience. In 44 chronic knee pain patients and 29 pain-free controls, we acquired CBF data (using arterial spin labelling) and T1-weighted images. Participants completed a series of questionnaires related to affective processes, and pressure and cuff algometry to assess pain sensitization. Two factor scores were extracted from these scores representing negative affect and pain sensitization, respectively. A spatial covariance principal components analysis of CBF identified five components that significantly discriminated chronic pain patients from controls, with the unified network achieving 0.83 discriminatory accuracy (area under the curve). In chronic knee pain, significant patterns of relative hypo-perfusion were evident in anterior regions of the default mode and salience network hubs, while hyperperfusion was seen in posterior default mode regions, the thalamus, and sensory regions. One component was positively correlated to the pain sensitization score (*r*=.43, *p*=.006), suggesting that this CBF pattern reflects the neural activity changes encoding pain sensitization. Here, we report the first chronic knee pain-related brain signature, pointing to a brain signature underpinning the central aspects of pain sensitisation.

## Introduction

Chronic musculoskeletal pain is a common public health, social and economic problem [32] with prevalence rates between 20 – 44% in the UK and US [11,13]. Unfortunately, as much as 40% of people living with chronic pain report unsatisfactory effect of treatment [7], highlighting a clear and urgent need to better understand chronic pain mechanisms for developing better treatments.

Recent work has focused on identifying specific neural signatures to explain the varied processes that contribute to the experience of physical pain [39], but relatively little progress has been made to unravel the neural basis of the chronic pain experience. Cerebral blood flow (CBF) as measured using arterial spin labeling (ASL) is particularly well-suited to study chronic pain, as it allows the capture of an absolute measure of non-evoked brain activity that underlies a “tonic” state such as ongoing spontaneous pain. To date, few CBF studies have investigated chronic pain, with even fewer studies in musculoskeletal (MSK) pain [10,19,21,40,41]. Observations of CBF changes in chronic MSK pain [19,40,41] are not consistent and are often limited by small sample sizes (N ≤ 17 pain patients). One recent study using a slightly larger sample (N=26 patients), did not observe any CBF differences between osteoarthritis patients and healthy controls, though did observe an association between ongoing pain and regional CBF in areas predominantly involved in emotional and fear regulation [10]. Most of these studies employ either voxel-wise or region-of-interest (ROI) methods which are known to lack the sensitivity to capture effects that may be more subtle and spatially distributed.

Multivariate analysis of CBF data provides a data-driven way to extract latent features of ongoing coordinated brain activity even from multiple parallel processes. This approach may allow the decoding of these processes and determine which markers distinguish patients with chronic pain from those without pain; a similar approach has been applied to distinguish patients with and without Parkinson’s disease [28]. Recently, a Gaussian Process classifier was applied to CBF data to discriminate pre- and post-surgical molar extraction intervention with just under 95% accuracy [29], demonstrating that certain CBF features are associated with *acute* severe pain in a clinical setting. To extend this further toward clinically relevant chronic pain, we aimed to identify novel discriminating markers of chronic pain in a population of largely community-dwelling people with mild-moderate knee pain, and explore how these features relate to specific facets of the chronic pain experience.

The purposes of this study were to use a multivariate approach (principal components analysis) to: i) discriminate patients with chronic knee pain from pain-free controls using non-evoked brain activity measures, and ii) determine whether these differences in CBF covariance patterns are related to affective or sensitized pain mechanisms in the pain syndrome. We hypothesized that the discriminatory components would comprise the classic pain connectome (insula, cingulate cortex) and extended pain networks including affective, sensory and arousal networks. Additionally, we hypothesized that these CBF patterns would differentially correlate with pain sensitization and affective pain phenotypes.

## Materials and methods

### Design and participants

This was a cross-sectional nested study within a larger knee OA multidimensional phenotyping study (INCOPE, Imaging Neural Correlates of Osteoarthritis Phenotypes) that will be reported in a future manuscript. For this CBF sub-study, the initial batch of the full INCOPE dataset comprising 54 patients with chronic knee pain and 33 healthy and pain-free participants was included (from a total dataset of 87 patients with chronic knee pain and 39 healthy and pain-free participants), allowing a reasonably matched group of patients and pain-free controls to test our multivariate approach on CBF data. Recruitment pathways were from a database of participants from the community via the East Midlands based Knee Pain and Related Health in the Community (KPIC) study cohort (Nottingham Research Ethics Committee 1, NREC reference 14/EM/0015; registered with ClinicalTrials.gov [NCT02098070]) [14,15] (N=53), primary care via general practitioner surgeries (N=27) within the Nottinghamshire region, or secondary care via the Sherwood Forest Hospitals NHS Foundation Trust orthopaedic referrals or poster advertisements at Nottingham City Hospital (N=7). All participants completed a set of questionnaires, quantitative sensory testing (QST), followed by an MRI session. The study was approved by the Nottingham Research Ethics Committee 2 (NREC reference: 10/H0408/115) and all participants provided written informed consent. Inclusion criteria for patients was self-reported diagnosis of knee osteoarthritis and/or reported chronic pain in the knee (i.e. pain for most of the day and pain for >14 days/month), and their knee pain was their most troublesome pain. Healthy participants reported no current or past history of knee pain (nor pain elsewhere). Participants were excluded if they had any history of stroke or had any current major neurological condition, psychosis, or had a contraindication to MRI (full list of inclusion and exclusion criteria is provided in the supplementary materials).

### Psychometric data

Participants all underwent psychometric assessments before the MRI scan session. Pain intensity on the day was taken in the hour prior to scanning using a numerical rating scale (NRS) ranging from 0 (no pain) to 100 (worst imaginable pain). Questionnaires included the Beck’s Depression Inventory II [6] (BDI-II), the Trait anxiety scale of the State-Trait Anxiety Inventory (STAI-T) [36] and the Pain Catastrophizing Scale [37], which was broken down into the subscales of helplessness, magnification and rumination. The BDI-II and the STAI-T were converted using Rasch conversion following the method of Lincoln et al [25], which recommended the BDI-II to be divided into two subscales (negative thoughts and negative behaviours) for measuring depression in patients with osteoarthritis.

### Quantitative Sensory Testing

All participants underwent quantitative sensory testing before the MRI scan session which consisted of pressure algometry (pressure pain thresholds, PPTs) using a handheld pressure algometer with a 1-cm^2^ probe and pressure was increased at a rate of approximately 30 kPa/s (Somedic AB, Sösdala, Sweden), and cuff pressure algometry using a computer-controlled cuff algometer (Cortex Technology and Aalborg University, Denmark). The pressure algometry assessed PPTs at two sites: the sternum and the most painful knee (or either knee in pain-free healthy controls). The cuff pressure algometry assessed pressure pain detection threshold (PDT), pressure pain tolerance threshold (PTT), temporal summation of pain (TS) and conditioned pain modulation (CPM) using the same method as described in previous studies [31,33,38]. Detailed methods of the quantitative sensory testing is provided in the supplementary materials.

### MRI Data acquisition

Participants underwent multimodal MRI at 3T (Discovery MR750, GE Healthcare) using a 32-channel head coil, as part of a larger phenotyping study (INCOPE) including arterial spin labelling [ASL] data and T1-weighted anatomical data used for image registration which are reported here for the CBF pattern sub-study. Cerebral blood flow was assessed using a pulsed-continuous ASL sequence with 3D spiral read-out (Tag/Control image pairs=72, Flip angle = 111°, TE=10.536ms, TR=4844ms, labelling duration=1450ms, post-labelling duration=2025ms, FOV=240mm, slice thickness=4mm, slice gap= 4mm, number of slices = 36, echo train length= 1, number of excitations = 3, matrix = 128 × 128, voxel resolution=1.875×1.875×4mm) [12]. ASL imaging also utilised background suppression and an M_0_ image for image quantification in line with current recommendations [2]. High resolution anatomical images were acquired in the sagittal plane using a fast spoiled gradient echo (FSPGR) sequence (TE/TR=3.164/8.132ms, TI=450ms, slice gap=1mm, field of view=256, matrix=256×256, flip angle=12°, voxel resolution=1mm^3^). T1-weighted images were acquired parallel to the AC-PC line whilst the bottom of the acquired ASL image was positioned just below the cerebellum to allow whole-brain CBF imaging.

### Image preprocessing

Each image was visually assessed for quality including artefacts induced by motion, and poor labelling defined by extremely low CBF values in the occipital regions (<20ml/100g/minute) relative to the rest of the cortex. Following quality control, there were 10 patients and 4 healthy control datasets excluded from the final analysis resulting in a total of 44 patients and 29 healthy and pain-free control subjects. Cerebral blood flow (CBF) maps (ml/100 g/min) were produced with the use of an automatic reconstruction script as reported in Zaharchuk et al [42]. Both the T1-weighted images and CBF maps were first skull-stripped using the brain extraction tool from FSL v5.0.11 (FMRIB Software Library). The CBF maps were then linearly registered to the T1-weighted images and then to MNI-space using FLIRT v6.0 (FMRIB’s Linear Image Registration Tool) [20]. The images were then spatially smoothed using a 5 mm full-width half-maximum Gaussian kernel and masked to exclude voxels that had a probability of less than 42% grey matter. This was chosen as it provided the most reliable CBF measures in an independent test-retest dataset (unpublished). These masked images were then used in the PCA analysis.

### Principal Components Analysis of CBF data

A voxel-based principal components analysis was used to generate patterns of spatial covariance in grey matter perfusion across the sample. We followed the method of Spetsieris et al [35] and Melzer et al [28] whereby the data was log-transformed and demeaned using the subject’s mean perfusion and the group mean, and then entered in a principal component analysis to calculate the eigenvectors and eigenvalues of the covariance. The spatial images of principal components with unit variance were generated using the same approach as it was proposed previously [28]. As a result, the first 16 components were selected, which explained 87.8% of the variance, with the first component captured the most variance, and the last component accounts the least. A backward stepwise binomial logistic regression was used to determine the components which successfully distinguished pain patients and pain-free controls (based on Akaike Information Criterion). These components were then linearly combined to create a unified network representing a chronic knee pain-related perfusion network and z-scored.

### Region of interest analysis

To validate the PCA findings, we also applied a region of interest analysis to determine whether the patterns of hyper- and hypo-perfusion were reflected in absolute CBF changes. The positive and negative loadings (thresholded at z>1.96 and z<-1.96 respectively) of the unified network and component 12 (the one component of 16 correlating with QST factor score) were masked onto each subject’s CBF map to extract mean CBF values. Age, sex and mean global CBF were regressed out of the absolute CBF data and all subsequent analyses were applied to the residuals. For the behavioural measures, both age and sex were regressed out of the data. Mean CBF of the unified network and component 12 were then compared between the two groups. In addition, the component 12 positive and negative loading CBF means for patients were correlated with the QST factor score to explore whether the relationship was driven by hyper- or hypoperfusion.

### Statistical Analyses

We performed the Kaiser-Meyer-Olkin test and Bartlett’s test of sphericity to ensure that a factor analysis was suitable for dimension reduction of both the psychometric and QST scores. This dimension reduction approach allows the integration of numerous variables gathered from psychometric questionnaire scores and QST scores to provide measures reflecting an overall measure of negative affect and sensitisation, respectively. Unrotated principal components analysis was used to extract a principal component for the psychometric scales and the QST scores using SPSS v25.0.0.1. These factor scores were used to relate to CBF measures. Linear regression was used to relate the network scores of the patient group with NRS pain severity score, affect factor score and QST factor score, correcting for age and sex. To test for the network reliability, a 500 permutation bootstrap estimation method was employed using the same approach as Melzer et al [28]. A z threshold of_1.96 was used to determine voxels that were considered reliable. For the ROI analyses of absolute CBF, age, sex and mean global CBF were regressed out of the data and all subsequent analyses were applied to the residuals. For the behavioural measures, both age and sex were regressed out of the data. Mean CBF comparisons between groups was carried out using an independents samples t-test. All analyses used an α level of *p*<.05.

A binomial generalised linear model was used to predict the labels of patients with chronic knee pain and healthy controls using each CBF component alone as well as the unified network and the area under the receiver operating characteristic curve (AUC) was analysed to assess the performance of the prediction. The area under the curve (AUC) of the receiver operating characteristic were obtained using Matlab 2018a for all the individual components and the unified network.

### Data Availability

Anonymised data may be made available upon request to the authors.

## Results

Demographics, psychometric and QST results in the final cohort of chronic knee pain patients and controls are provided in Table 1. Table of medications are included in the supplementary materials.

**Table 1.** Demographic and clinical data of participants

| Data | Knee pain patients | Healthy controls | p value |
| --- | --- | --- | --- |
| N | 44 | 29 | – |
| Mean age | 62.8 (8.6) | 64.4 (11.1) | 0.49 |
| Sex (male/female) | 22/22 | 18/11 | 0.31 |
| Laterality of affected knee (L/R) | 22/22 | – | – |
| Mean educational scores | 5<sup>a</sup> | 3<sup>a</sup> | **0.02** |
| Pain duration (months) | 119.7 (121.9) | – | – |
| NRS pain 0 – 100 on the day | 36.3 (29.4)<sup>a</sup> | – | – |
| PainDETECT (Rasch converted) | -0.6 (0.7)<sup>a</sup> | – | – |
| BDI-II negative thoughts subscale (Rasch converted) | 2.9 (3.0) | 1.7 (2.2) | 0.07 |
| BDI-II negative behaviours subscale (Rasch converted) | 9.4 (3.3) | 5.3 (3.2) | **<0.001** |
| STAI-T (Rasch converted) | -1.3 (1.5) | -1.8 (0.9) | 0.124 |
| PCS | 18.4 (14.4)<sup>a</sup> | 9.2 (7.4) | **<0.01** |
| PCS: helplessness | 8.5 (6.3)<sup>a</sup> | 3.2 (3.1) | **<0.01** |
| PCS: magnification | 3.4 (3.4)<sup>a</sup> | 1.9 (1.6) | **0.04** |
| PCS: rumination | 6.8 (5.2)<sup>a</sup> | 4.1 (3.6) | **0.02** |
| PPT sternum [kPa] | 260.5 (162.3)<sup>a</sup> | 300.4 (161.7)<sup>a</sup> | 0.31 |
| PPT knee [kPa] | 308.7 (202.5)<sup>a</sup> | 398.9 (221.5)<sup>a</sup> | 0.08 |
| PDT affected leg [kPa] | 17.5 (6.81)<sup>a</sup> | 21.3 (8.0)<sup>b</sup> | **0.04** |
| PTT affected leg [kPa] | 32.8 (13.3)<sup>a</sup> | 39.4 (14.8)<sup>b</sup> | 0.06 |
| PDT unaffected leg [kPa] | 17.2 (6.9)<sup>a</sup> | 19.6 (9.4)<sup>c</sup> | 0.23 |
| PTT unaffected leg [kPa] | 30.6 (12.1)<sup>a</sup> | 35.7 (13.9)<sup>c</sup> | 0.11 |
| TS [VAS points] | 1.0 (1.3)<sup>b</sup> | 0.6 (1.0)<sup>b</sup> | 0.16 |
| CPM [kPa] | 4.83 (8.36)<sup>b</sup> | 9.5 (10.3)<sup>c</sup> | **0.04** |
| Affect factor score | 0.0 (1.1)<sup>a</sup> | -0.5 (0.5) | **0.001** |
| QST factor score | 0.3 (1)<sup>b</sup> | 0.6 (1.0)<sup>c</sup> | **0.03** |

Values displayed are means and standard deviations (in parentheses). Education is scored with 1 with highest level of qualification to 8 with no formal education. PainDETECT Rasch conversion according to Lincoln et al (2017). BDI-II – Beck's Depression Index, NRS – numerical rating scale, STAI-T – Trait Anxiety, PCS – Pain Catastrophizing Scale, PPT – Pressure Pain Threshold, PDT – Pain Detection Threshold, PTL – Pain Tolerance Threshold, TS – Temporal Summation, CPM – Conditioned Pain Modulation <sup>a</sup>one subject score missing; <sup>b</sup>two subject scores missing; <sup>c</sup>three subject scores missing. BOLD indicates significant group differences (uncorrected for multiple comparisons).

### Psychometric data

Between groups comparisons showed significant differences for BDI-II negative behaviours subscale, PCS and all PCS subscales (Table 1). The Kaiser-Meyer-Olkin measure (0.81) and significant Bartlett’s test of sphericity (*p*<.0001) indicated that a factor analysis was suitable for dimension reduction of the BDI-II subscales, STAI-T and the PCS subscale scores. The extracted principal component explained 68.7% of the variance. This component loaded positively on all scores with maximum loading for PCS helplessness, where higher factor scores indicated more negative affective characteristics (Table 2).

**Table 2.** Component loading scores for the affective measures

| **Affect measures** | **Component 1** |
| --- | --- |
| Trait anxiety (STAI-T) | .81 |
| PCS helplessness | .91 |
| PCS magnification | .87 |
| PCS Rumination | .83 |
| BDI-II negative thoughts | .80 |
| BDI-II negative behaviours | .75 |
| BDI – Beck's Depression Index, STAI-T – Trait Anxiety, PCS – Pain Catastrophizing Scale |  |

### QST data

Between groups comparisons revealed a significantly lower PDT of the affected leg and reduced CPM in patients (Table 1). There was also a trend toward greater sensitivity in patients for the PPT of the knee and PTT of the affected leg (*p*<0.1, Table 1). A total of 8 scores were derived from the QST (PPT of sternum and knee, PDT and PTT of each leg, TS and CPM). The Kaiser-Meyer-Olkin test (0.65) and Bartlett’s test of sphericity (*p*<.0001) indicated suitability of a factor analysis for dimension reduction of the QST scores. The extracted component explained 54.5% of the total variance which loaded positively and highly on all measures indicating greater pain sensitivity (maximum ipsilateral pain tolerance threshold) except for the TS, where high scores indicate greater sensitivity resulting in negative loading albeit of much smaller extent (Table 3).

**Table 3.** Component loading scores for the QST measures

| **QST measures** | **Component 1** |
| --- | --- |
| PPT sternum | .77 |
| PPT knee | .68 |
| TS | -.16 |
| CPM | .75 |
| PDT affected leg | .82 |
| PTT affected leg | .89 |
| PDT unaffected leg | .77 |
| PTT unaffected leg | .81 |
| PPT – Pressure Pain Threshold, PDT – Pain Detection Threshold, PTT – Pain Tolerance Threshold, TS – Temporal Summation, CPM – Conditioned Pain Modulation |  |

### Principal components of CBF data

The PCA extracted 16 components that were used for predicting group classification. Components 2, 6, 8, 12 and 13 significantly classified knee pain patients from controls. These components explained 19.9%, 2.2%, 1.3%, 1% and 0.9% of the variance respectively and were considered to be knee pain-related components. The area under the curve (AUC) of the unified network was 0.83 while the five included individual components yielded 0.62-0.64 classification power. The unified network achieved 82% specificity and 76% sensitivity. The unified component and component 12 are illustrated in Figure 1a and 1b respectively, with details of cluster regions (clusters > 20 voxels) listed in Table 4.

**Table 4.** Cluster maxima of the unified of components that significantly classified knee pain patients from controls.

| Region | Cluster size<br>(voxels) | MNI coordinates |  |  | Z value |
| --- | --- | --- | --- | --- | --- |
|  |  | X | Y | Z |  |
| *Hyperperfusion in chronic knee pain* |  |  |  |  |  |
| Lateral occipital, cuneus, precuneus, posterior cingulate cortex | 3320 | -14 | -84 | 40 | 4.03 |
| Cerebellum | 1907 | 8 | -88 | -32 | 3.84 |
| Fusiform gyrus, inferior temporal gyrus | 349 | -42 | -26 | -22 | 2.99 |
| Inferior temporal gyrus, lateral occipital cortex | 252 | 54 | -62 | -12 | 2.85 |
| Lingual gyrus, parahippocampal gyrus | 131 | -14 | -42 | -10 | 2.4 |
| Lateral occipital cortex | 124 | -40 | -76 | 18 | 3.11 |
| Cerebellum | 83 | -26 | -52 | -56 | 3.1 |
| Cerebellum | 52 | -6 | -50 | -60 | 2.86 |
| Brain stem | 46 | -4 | -44 | -62 | 3.86 |
| Temporal pole | 43 | -48 | 22 | -24 | 2.8 |
| Thalamus | 41 | 4 | -18 | 10 | 2.4 |
| Inferior temporal gyrus | 39 | 48 | -6 | -46 | 2.71 |
| Lingual gyrus | 38 | 16 | -68 | -2 | 2.54 |
| Parahippocampal gyrus | 36 | -26 | -22 | -20 | 2.49 |
| Frontal pole | 28 | -12 | 66 | 24 | 2.18 |
| Lateral occipital cortex, fusiform gyrus | 23 | -44 | -74 | -16 | 2.53 |
| Frontal pole | 22 | -6 | 70 | 8 | 2.26 |
| Parahippocampal gyrus, fusiform gyrus | 21 | 26 | -10 | -38 | 2.52 |
| Caudate | 20 | -16 | -10 | 24 | 2.49 |
| *Hypoperfusion in chronic knee pain* |  |  |  |  |  |
| Supramarginal gyrus, postcentral gyrus, parietal operculum | 1339 | -58 | -20 | 28 | 3.42 |
| Frontal pole | 1004 | 30 | 54 | 10 | 3.52 |
| Cerebellum | 633 | 28 | -38 | -50 | 4.68 |
| Heschl's gyrus, planum polare, insula | 537 | -44 | -16 | 6 | 3.35 |
| Anterior cingulate cortex | 533 | -2 | 42 | 10 | 3.07 |
| Orbitofrontal cortex | 441 | 0 | 30 | -20 | 3.23 |
| Orbitofrontal cortex, anterior insula | 392 | 36 | 24 | -8 | 3.84 |
| Opercular cortex | 241 | 42 | -12 | 18 | 2.89 |
| Cerebellum | 219 | -48 | -58 | -50 | 3.81 |
| Postcentral gyrus, supramarginal gyrus | 208 | 52 | -26 | 40 | 2.89 |
| Frontal pole | 185 | -32 | 52 | 6 | 2.97 |
| Mid-cingulate gyrus | 121 | 0 | -4 | 36 | 2.5 |
| Orbitofrontal cortex, inferior frontal gyrus, anterior insula | 110 | -34 | 30 | -2 | 2.89 |
| Postcentral gyrus, supramarginal gyrus | 101 | 38 | -32 | 42 | 3.14 |
| Precentral gyrus, inferior frontal gyrus | 89 | 54 | 10 | 32 | 3.23 |
| Lateral occipital cortex, angular gyrus | 74 | 42 | -60 | 56 | 2.83 |
| Lateral occipital cortex, superior parietal cortex | 67 | -24 | -58 | 68 | 2.37 |
| Middle frontal gyrus, inferior frontal gyrus | 63 | 42 | 22 | 30 | 3.14 |
| Anterior cingulate cortex | 62 | 12 | 40 | 14 | 2.62 |
| Cerebellum | 54 | -28 | -82 | -44 | 2.56 |
| Inferior frontal gyrus, precentral gyrus | 51 | 56 | 10 | 12 | 3.02 |
| Inferior temporal gyrus, middle temporal gyrus | 35 | -56 | -10 | -32 | 2.6 |
| Superior temporal gyrus, planum temporale, Heschl's gyrus | 34 | -58 | -4 | -2 | 2.57 |
| Superior frontal gyrus | 34 | 12 | 10 | 62 | 2.93 |
| Orbitofrontal cortex | 31 | -20 | 30 | -22 | 2.69 |
| Middle frontal gyrus | 30 | 40 | 10 | 48 | 2.82 |
| Planum temporale, insula | 26 | -46 | 2 | -12 | 2.43 |
| Frontal pole | 23 | -24 | 44 | 20 | 2.43 |
| Orbitofrontal cortex | 22 | 22 | 12 | -16 | 2.38 |
| Angular gyrus, supramarginal gyrus | 22 | -50 | -52 | 28 | 2.52 |
| Superior frontal gyrus, supplementary motor cortex | 21 | 10 | 10 | 58 | 2.91 |

\*Only clusters >20 voxels in size are listed.

**Figure 1.**
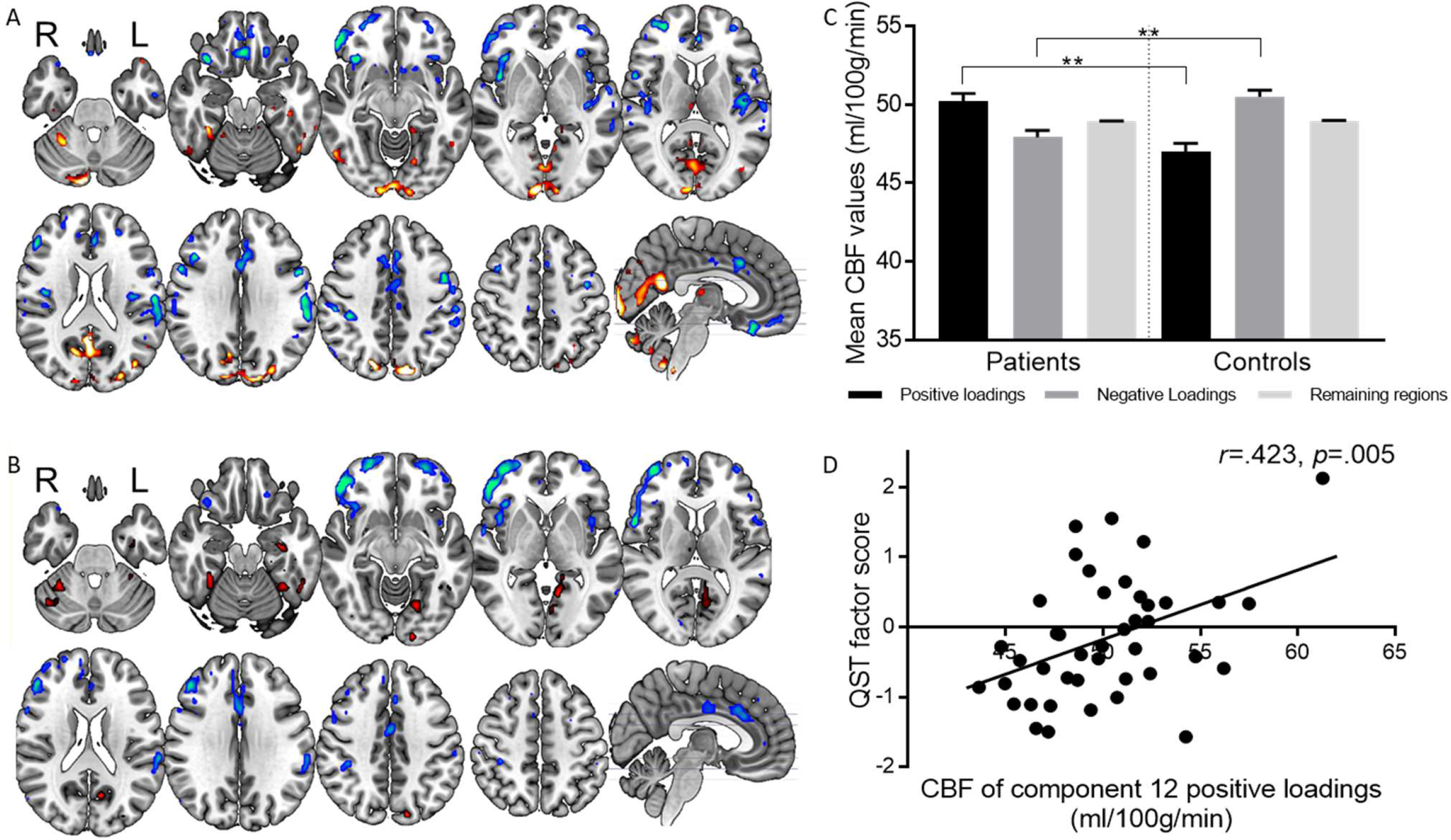
Components discriminating people with chronic pain and healthy controls and their association with pain sensitisation. A) The unified knee pain-related network (combination of five components) that classified patients and controls. Clusters in hot colours are regions with positive loadings, clusters in cold colours are regions with negative loadings. B) Component 12 from the PCA that classified patients from controls. This component was also significantly correlated with the QST factor score. Clusters in hot colours are regions with positive loadings, clusters in cold colours are regions with negative loadings. C) The mean absolute CBF values extracted from the unified network within regions with positive, negative loadings and the remaining regions outside of the unified component, in knee pain patients and controls (plotted with standard error of the mean). D) Mean CBF of the regions with positive loadings of component 12 (red clusters in B) correlated with the QST factor score.

Of the five discriminatory components, only component 12 was significantly associated with the QST factor score (*r*=.43, *p*=.006). The affect factor score was not significantly correlated with any of the knee pain-related components. Correlation between the discriminating components and the individual QST and affect measures can be found in the supplementary materials. Pain severity did not significantly correlate with any of the five discriminatory components.

### Region of interest analysis

#### Unified network

Mean CBF was significantly higher in patients with knee pain (M=50.23, SEM=0.48) compared with healthy controls (M=47.01, SEM=0.54) within the knee pain-related perfusion network with positive loadings (t(71)=4.38, *p*<.001, Fig. 1c), and significantly lower CBF (knee pain patients: M=47.93, SEM=0.43; pain-free controls: M=50.49, SEM=0.43) within the regions with negative loadings (t(71)=-4.02, *p*<.001, Fig. 1c), confirming that the positive and negative loadings reflect hyper- and hypoperfusion respectively. Conversely, mean CBF outside of the pain-related network (i.e. grey matter regions that did not reach the threshold for positive or negative loadings) did not differ between chronic knee pain and pain-free participants (knee pain patients: M=48.94, SEM=0.02; pain-free controls: M=48.97, SEM=0.02).

#### Component 12

Clusters from component 12 with positive and negative factor loading showed significant hyperperfusion (t(71)=3.26, *p*=.002), and hypoperfusion (t(71)=-2.36, *p*=.02) in pain patients versus controls. Within the patient group absolute CBF averaged over the networks of component 12 showed a significant positive association with the QST factor for the positive regions (*r*=.423, *p*=.005) but no significant association for the negative regions (*r*=-.224, *ns*). These results are illustrated in Figure 1d. A closer inspection of this CBF correlation was carried out to explore the correlation with individual QST measures which showed an association with all measures except for TS (these results are provided in the supplementary materials).

In addition, as the chronic pain signature did not include the PAG as expected [9,19,22], we undertook a regional PAG posthoc analysis to mitigate against the risk that the PCA dimensionality reduction may fail to detect small clusters forming local networks. As shown in the supplementary results, mean PAG CBF suggested hyperperfusion in patients compared with controls.

## Discussion

Using PCA of CBF data, the first clinical pain signature was derived from non-evoked brain activity in chronic knee pain patients compared with pain-free controls. The discriminating knee pain-related network revealed a covariance pattern of hypo-/hyperperfusion within extended pain connectome regions. A component of the hyperperfusion network was related to the degree of pain sensitisation pointing to a brain signature underpinning the central aspects of pain sensitisation.

### Knee-pain related network and its link to sensitisation measures

The ability of CBF covariance maps to clearly discriminate between chronic pain and pain-free groups highlights the dual advantage of combining a multivariate pattern approach with a quantitative physiological measure of brain activity as previously exploited for acute pain [29]. There is however, evidence demonstrating spatial correlation between network hubs measured via fMRI and CBF [24] allowing comparisons across modalities, albeit with caution. The knee pain-related network corresponds well with regions considered part of the pain connectome from resting-state fMRI, with a distinct pattern of increased perfusion in posterior DMN regions and reduced perfusion in salience network regions. Previous studies commonly report dysfunctions of regions comprising both the DMN and salience network in various chronic pain conditions and across a range of imaging modalities [3,4,8,19,23,27]. In one study using CBF data to discriminate pre- and post-surgical third molar extraction, the multivariate pattern of perfusion classifying post-surgical painful state [29] is in striking contrast to the current unified component classifying chronic knee pain patients. They showed increased CBF post-surgery in the thalamus, salience network, secondary somatosensory, and anterior cingulate cortices, with decreased CBF in visual and parietal cortices. This dissociative pattern between our knee pain patients with mild ongoing pain and the post-surgery patients with severe acute pain from O’Muircheartaigh et al.’s study, thus largely excludes upregulated nociception as underlying mechanism, and conversely suggests a maladaptive neuroplasticity process linked to pain chronification.

The observed CBF pattern of chronic pain showed however an overlap with the acute postsurgical pain CBF signature reported in O’Muircheartaigh et al., with both displaying increased CBF of the thalamus. It is tempting to speculate that persistent thalamic activation may be a key process in driving pain chronification. In fact, thalamic volume changes have been observed in patients with chronic hip OA pain, moreover, this effect is normalised following successful arthroplasty intervention [17]. In our data, the dorsomedial nucleus was specifically observed to be hyperperfused in patients and this was seen across the unified components map as well as the component that showed a correlation with QST measures. This nucleus is part of the medial pain pathway that is responsible for the affective-motivational elements of pain perception, and moreover has extensive connections with the cortex including somatosensory and limbic regions [1], akin to those regions we observed to be hypoperfused in patients with chronic knee pain. The association between one of the discriminating components with the QST score aligns well with the notion that aberrancies within this network are more pronounced in those who are more centrally sensitised to pain. The continuous nociceptive barrage may result in a sensitised state of the thalamic circuitry that links to numerous areas of higher-order cognitive processes [1]. This cascading effect of the overactive thalamus may explain the lack of deactivation of posterior DMN regions in our patients and perhaps extend to affect other sensory (visual) regions. The reverse pattern observed in the aforementioned acute post-surgical pain study [29] may be a demonstration of the normal function of this thalamic circuitry to effectively up- and down-regulate salience and default mode regions respectively during an acute pain experience. In contrast, in chronic knee pain patients, continuous peripheral input may have resulted in a breakdown of this system leading to dysfunctional bottom-up processes and an abnormal interaction between salience and default mode networks. This constant peripheral pain drive has previously been proposed to maintain sensitization of central pain pathways in MSK pain [16].

Kucyi et al. [22] found that spontaneous mind-wandering away from pain resulted in increases in DMN activity with greater salience network activity when attending to pain. This is consistent with the present findings of increased and decreased perfusion in the DMN and salience regions respectively in patients who had lower sensitivity to pain. The higher activity of the posterior cingulate, precuneus and cuneus regions in less sensitised patients may allow these patients being better able to assess and appropriately orient attention toward and/or away from pain. We speculate that the greater effectiveness of this system may have enabled patients to become better adapted to cope with frequent nociceptive input, and thus preventing central sensitisation. The deactivation of regions such as the insula and ACC may be reflective of hypoarousal and a lesser ability for interoception. There may be a complex interplay between the salience and default mode networks that results in an impaired capacity to effectively disengage their attention to pain. Although purely speculative, this difference in the ability to attend to or disengage from pain may determine the reorganisation of one’s brain circuitry during the chronification and central sensitisation process.

### Changes in the periaqueductal grey in chronic knee pain

It is however, surprising that the PAG was not evident as a pain-relevant region in our cohort given previous reports of PAG changes in chronic pain [9,18,19,22,34] and its well-documented involvement in the antinociceptive system for inhibition of pain [5,26,30]. One might expect this region to robustly differentiate chronic knee pain patients from pain-free controls, and furthermore be related to sensitisation of central pain mechanisms, particularly if our theory above regarding the attending toward/away from pain is true. However, the spatial resolution that is feasible with ASL does not allow the segregation of the subcomponents of the PAG and therefore the signal measured from the PAG will be an average across both the ascending and descending pain pathways which are known to have distinct connections and mechanisms underlying the pain experience [26]. Furthermore, small clusters are less likely to be detected using a PCA approach due to the thresholds applied for explained variance. A posthoc exploratory analysis of the PAG in our dataset revealed that, similarly to a previous report [19], there is indeed increased perfusion in patients with chronic knee pain (supplementary figure 1), suggesting that although there are PAG alterations in chronic knee pain, the PAG perfusion pattern may not contribute to the major CBF networks derived via the PCA approach across the full dataset. It would be of interest to further investigate in the same individuals whether there is a link between CBF increases of the PAG (as seen in the current study) and altered connectivity of the PAG with default mode regions (as seen in [22]).

### Negative affect in chronic knee pain

The lack of association between the affective score and CBF is not unexpected. Although our patient cohort was significantly higher scoring for affective elements, they would still be considered to be within the healthy range for levels of depression and anxiety. Therefore, the present dataset may lack the variation in affective features to be able to effectively test our hypothesis. We also suggest that the PCA approach be applied to a patient group (ideally with high variation of affective measures) to identify components that differentiate patients with high and low negative affective features in order to better deconstruct the effect of psychological aspects contributing to chronic pain. In addition, the natural next step is to validate this model using an independent dataset to test the predictive confidence and generalisability.

### Limitations

A limitation of our study is that a proportion of our patients had taken opioid medication and/or antidepressants which could have affected CBF especially within the salience and default mode regions. However, comparison of the mean CBF values of both the positive and negative loadings of the unified component between those on opioid or antidepressant medication and those who were not, showed no significant difference (see supplementary materials for detailed results). Moreover, visual inspection of the correlation between the QST factor score and both the component 12 network score and the mean CBF of component 12 positive loadings did not show a systematic pattern to suggest there was a strong impact of medication on the current findings (colour-coded correlation plot of Fig 1d is available in the supplementary materials).

## Conclusions

This study successfully delineated patients with chronic knee pain from controls using a PCA approach on CBF data, and the discrimination was related to the degree of pain sensitisation as measured via comprehensive quantitative sensory testing methods. The patterns of CBF alterations indicate greater blood flow changes in primarily the salience and default mode network regions commonly implicated in chronic pain, and these were further heightened in those with greater central sensitisation. This pattern was unrelated to the pain severity on the day and largely opposite to the reported CBF signature of acute postsurgical pain further pointing to a neuroplastic, and also a potentially non-nociceptive origin. We demonstrate that task-free CBF assessment contains important covariance features that may signify the neural mechanisms of pain chronification and pain sensitisation.

## Data Availability

Anonymised data may be made available upon request to the authors.

## Acknowledgements

The authors are also indebted to colleagues who enabled recruitment (Nadia Frowd, Dr Bonnie Millar, Marya Habib, Louise Borg and Debbie Wilson) and scanned participants (Andrew Cooper). The authors thank Versus Arthritis (Grant 20777), Parkinson’s UK (Grant J□1204), NIHR Nottingham Biomedical Research Centre and Michael J Fox Foundation (Grant 11473) for providing the financial support for this study. Center for Neuroplasticity and Pain (CNAP) is supported by the Danish National Research Foundation (DNRF121). All authors declare no conflict of interest.

